# Domain-Specific Effects of Transcranial Direct Current Stimulation in Chronic Lower Back Pain: A Bayesian Internal Meta-Analysis

**DOI:** 10.1101/2024.11.25.24317873

**Authors:** Emily J. Corti, Natalie Gasson, An T. Nguyen, Andrea M. Loftus, Welber Marinovic

## Abstract

Chronic pain is associated with deficits in cognitive functioning and increased pain intensity. Research investigating the potential for transcranial direct current stimulation (tDCS) to improve cognitive functioning, pain experience, and psychological health in individuals with chronic lower back pain (CLBP) yield mixed results. The present randomised, placebo-controlled study examined the impact of anodal-tDCS over left-dorsolateral prefrontal cortex (left-DLPFC) on cognitive functioning, pain experience, and psychological health in those with CLBP. Thirty participants with CLBP (Mage = 57.47 years, SDage = 14.28 years) received 20-minutes of sham or anodal tDCS, twice weekly, for 4 weeks. Twenty-six outcomes across five functional domains were assessed at baseline and post-intervention. Bayesian multilevel models estimated intervention effects for each outcome, and an internal meta-analysis synthesised domain-level effects. Results revealed robust cognitive enhancement for Executive Function/Attention (pooled effect 0.21 SD, 95% highest posterior density [HPD] [0.03, 0.40]), with the largest effect observed for Letter Number Sequencing (0.56 SD, 95% HPD [0.09, 1.04]). Pain & Physical Function showed borderline effects (0.18 SD, 95% HPD [-0.03, 0.38]), while Mood & Wellbeing showed negligible effects (0.03 SD, 95% HPD [-0.15, 0.22]). These findings demonstrate domain-specific effects, with selective cognitive improvements but unclear analgesic effects and no mood benefits. Anodal tDCS may improve cognitive deficits in CLBP but should not be considered a standalone intervention for pain or mood. Future research should examine alternative cortical targets for pain relief and potential synergistic effects with psychological interventions.

## Introduction

Chronic lower back pain (CLBP) is a multidimensional condition characterised by alterations in cognition, psychological health, and quality of life. However, current recommended treatments typically focus only on pain and physical symptoms of the condition. These treatments usually result in small to modest improvements in the short-term but are ineffective for long-term pain management (Chou et al., 2017). It has been suggested that management of CLBP should encompass a biopsychosocial approach that aims to improve not only pain experience, but also functionality (cognition and disability) and psychological wellbeing (mood and quality of life; Cuomo et al., 2021).

The prefrontal cortex plays a key role in pain modulation (Lorenz et al., 2003). Functional imaging studies have revealed that the dorsolateral prefrontal cortex (DLPFC) is involved in both pain modulation (Brighina et al., 2011) and the cognitive evaluation of pain (Melzack, 2001; Mylius et al., 2006). Research suggests that the processing of pain engages significant neural resources in the DLPFC. Consequently, there are fewer neural resources available for other cognitive functions of the DLPFC, such as planning and working memory (Berryman et al., 2013; Seminowicz & Davis, 2007). It has also been suggested that decreased cortical inhibition extends the duration of activation in the DLPFC during pain processing. During this time, the DLPFC is unable to shift attention away from the pain and reallocate those resources to other cognitive tasks (Berryman et al., 2013). Attending to pain is therefore highly cognitively demanding and leaves little resources available in the DLPFC for normal functioning (Moseley, 2003; Smith & Ayres, 2014)

Research has provided strong evidence of cognitive deficits in people with chronic pain (Higgins et al., 2018). However, there are discrepancies regarding which cognitive domains are affected and whether this differs between pain conditions. Compared to controls, people with chronic pain perform poorly on general measures of cognition (Oosterman et al., 2010; Weiner et al., 2006) and the prevalence of significant global deficit (≤ 24 on the Mini Mental State Examination) is higher in the chronic pain population compared to the general population (Povedano et al., 2007; Rodriguez-Andreu et al., 2009). Deficits in attention have been reported in multiple chronic pain conditions, including CLBP (Dick & Rashiq, 2007) and Fibromyalgia (Dick et al., 2002). A systematic and meta-analytic review reported working memory deficits in multiple chronic pain conditions, including CLBP and non-specific CLBP (Berryman et al., 2013). Specific differences between people with chronic pain and healthy controls were reported in verbal working memory, non-verbal working memory, and attention (Berryman et al., 2013). However, no differences were identified in spatial working memory between people with chronic pain and healthy controls (Berryman et al., 2013). In comparison, a review by Moriarty et al. (2011) reported that people with chronic pain performed poorly on measures of verbal and spatial working memory. These findings were consistent with research in CLBP, that reported deficits in verbal, visual, and spatial memory when compared to pain-free controls (Jorge et al., 2009).

While the specific impairment may differ between pain conditions, cognitive impairment is a common feature of chronic pain that significantly impacts treatment outcomes. Cognitive impairment in people with chronic pain can impact upon a person’s planning ability, mental flexibility, and decision making (Apkarian et al., 2004; Berryman et al., 2013; Moriarty et al., 2011), rendering cognitive-behavioural rehabilitation strategies ineffective (Smeets et al., 2006). Poor cognitive performance is associated with disengagement from daily activities, reduced treatment adherence, and poor quality of life in people with chronic pain (Hoy et al., 2014, Roth et al., 2005). As such, effective management of chronic pain needs to encompass strategies to improve cognitive function.

Transcranial Direct Current Stimulation (tDCS) is a non-invasive brain stimulation technique that delivers low intensity electrical currents to modulate neural activity (Nitsche et al., 2008). Anodal-tDCS (a-tDCS) over left-DLPFC improves cognitive function in numerous conditions, such as neurodegenerative disease, and healthy ageing (Boggio et al., 2006; Boggio et al., 2009; Coffman et al., 2014). Research in healthy individuals has found that tDCS over DLPFC improves cognitive functioning (Coffman et al., 2014). Participants who experience problems with their memory show improved cognitive functioning across multiple cognitive domains following a-tDCS over DLPFC (Hansen, 2012). Research in neurodegenerative diseases has also reported improvements in cognitive functioning following a-tDCS over DLPFC (Boggio et al., 2006; Boggio et al., 2009; Hansen, 2012). Participants with Parkinson’s disease improved on a working memory task following a single session of a-tDCS over DLPFC (Boggio et al., 2006). Participants with Alzheimer’s improved on a visual recognition memory task following a single session of a-tDCS over DLPFC (Boggio et al., 2009). While there is substantial evidence for the use of tDCS over left-DLPFC to improve cognition in clinical and healthy ageing populations, few studies have investigated the impact of tDCS on cognitive functioning in chronic pain.

Research in fibromyalgia indicated a trend towards improvement in global cognition following five daily sessions of a-tDCS over DLPFC (Fregni et al., 2006). Improvements were also shown on an attention and working memory task, and simple reaction time task (Fregni et al., 2006). Silva et al. (2017) reported improvements in selective attention (orientating and executive) in people with fibromyalgia following a single session of a-tDCS over DLPFC. It is believed that a-tDCS induces long-term potentiation-like plasticity mediated by upregulating N-methyl-d-aspartate (NMDA) and GABA receptor activity. These receptors play a key role in maintaining optimal cognitive function (Ciampi de Andrade et al., 2014). In chronic pain, a-tDCS over left-DLPFC is thought to inhibit the allocation of maladaptive cognitive and attentional resources to pain, such that people disengage their attention from their pain and assign those resources to other cognitive functions (Berryman et al., 2013; Smith & Ayres, 2014). For those with chronic pain, the inhibition of maladaptive cognitive evaluations of pain may help to alleviate the pain and improve cognitive functioning. However, the impact of tDCS over DLPFC on cognition has not yet been examined in CLBP.

Limited studies have shown tDCS over DLPFC improves cognitive functioning in certain chronic pain conditions (Fregni et al., 2006; Silva et al., 2017). A-tDCS over DLPFC reduces pain levels in some forms of chronic pain (Brietzke et al., 2020; To et al., 2017). It is unclear whether a-tDCS over left-DLPFC can improve cognitive functioning in people with CLBP and, if so, whether there is a corresponding improvement in pain-related outcomes and psychological health. For the present study, it was proposed that 2x weekly, 1.5mA a-tDCS over left-DLPFC for 4 weeks would improve cognitive function in people with CLBP, whereby participants in the a-tDCS group would demonstrate an increase in cognitive performance compared to a sham-tDCS group (s-tDCS). It was also proposed that the a-tDCS group would demonstrate a reduction in pain-related outcomes (pain intensity, disability, and pain catastrophising), compared to the s-tDCS group. Finally, it was proposed that the a-tDCS group would demonstrate an improvement in psychological outcomes (depression, stress, anxiety, and quality of life) compared to the s-tDCS group.

## Methods

### Participants

Participants were recruited to participate in a 5-week intervention study. This study was approved by Curtin University Human Research Ethics Committee (HR17/2015) and all research was conducted in accordance with the Declaration of Helsinki. All participants provided written, informed consent. Inclusion in the study required a formal diagnosis of CLBP by a qualified health professional (General Practitioner or Physiotherapist) of at least 6 months (see Table 1 for demographics and pain related information). Individuals’ eligibility was assessed against a tDCS screening questionnaire (Nitsche et al., 2008) and individuals were screened for cognitive status using the Telephone Interview for Cognitive Status – 30 (TICS-30; score ≥ 18 for inclusion). Thirty-one participants met the inclusion criteria. One participant withdrew from the study prior to the first session of the intervention and was not included in the analysis. Thirty participants completed the intervention (see Figure 1).

**Figure 1.**
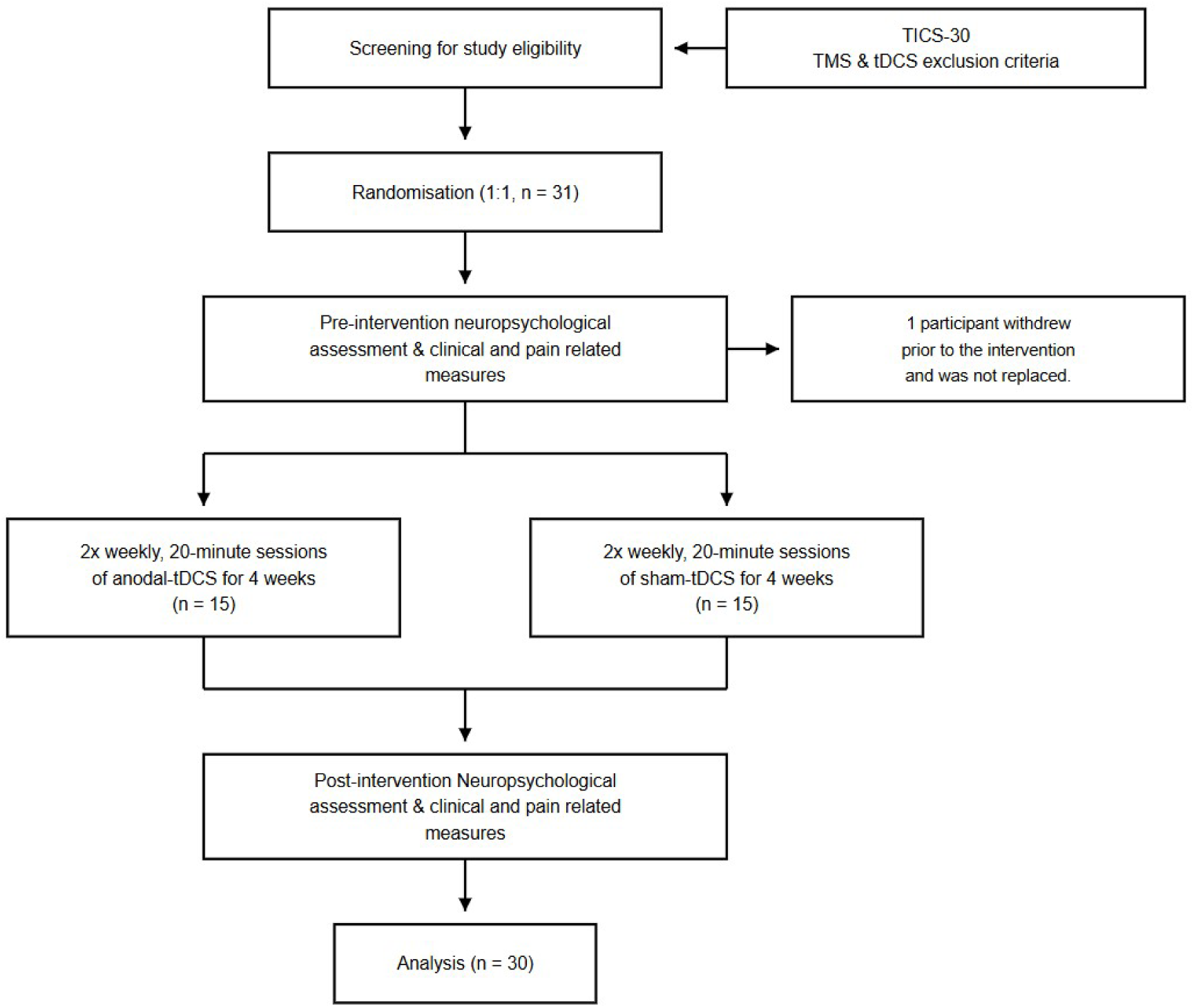
Flow diagram of the progress of the trial for anodal and sham transcranial direct current stimulation (tDCS) groups.

**Table 1.**
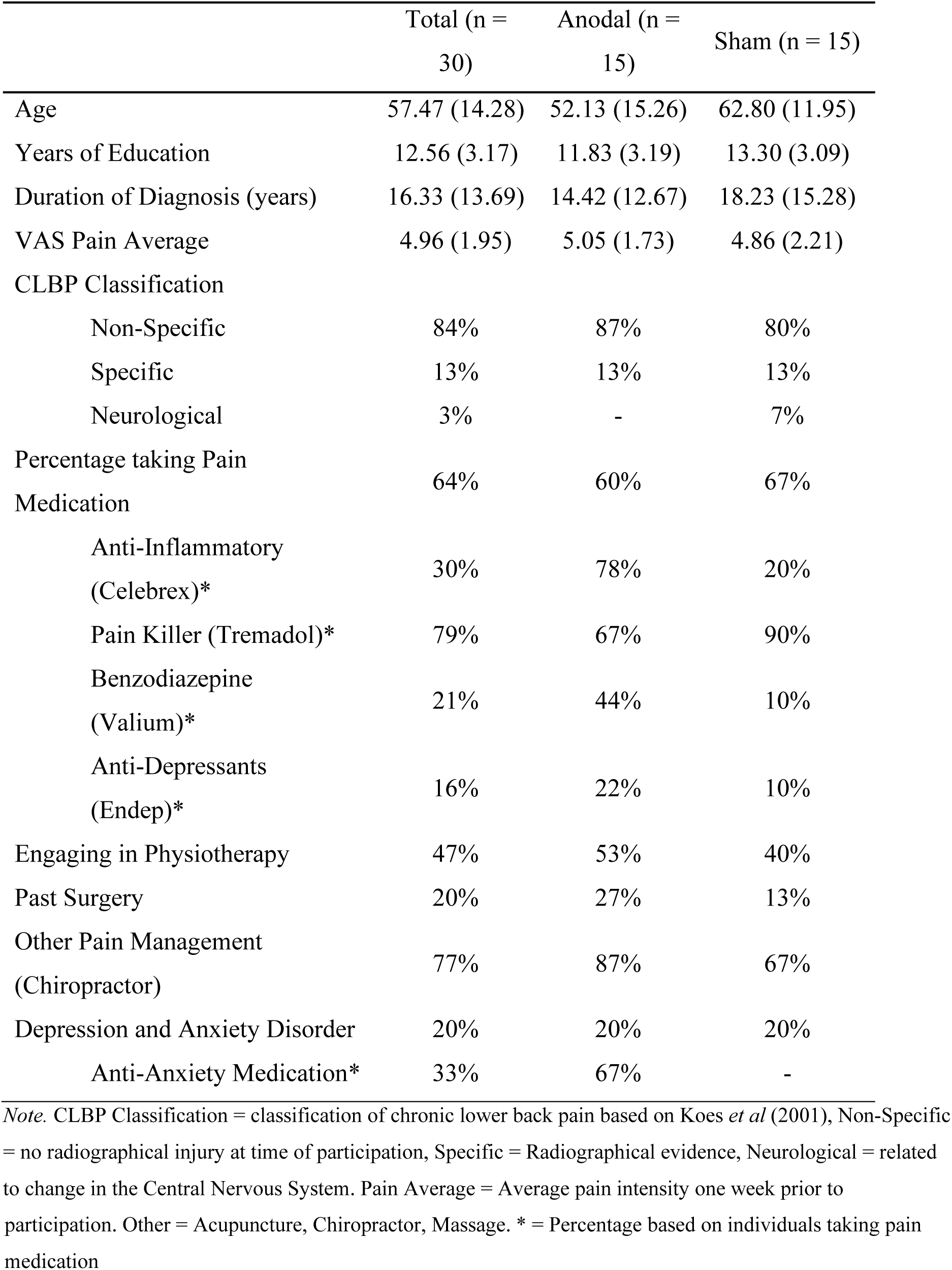
Baseline demographics and pain-related information by intervention group.

|  | Total (n = 30) | Anodal (n = 15) | Sham (n = 15) |
| --- | --- | --- | --- |
| Age | 57.47 (14.28) | 52.13 (15.26) | 62.80 (11.95) |
| Years of Education | 12.56 (3.17) | 11.83 (3.19) | 13.30 (3.09) |
| Duration of Diagnosis (years) | 16.33 (13.69) | 14.42 (12.67) | 18.23 (15.28) |
| VAS Pain Average | 4.96 (1.95) | 5.05 (1.73) | 4.86 (2.21) |
| CLBP Classification |  |  |  |
| Non-Specific | 84% | 87% | 80% |
| Specific | 13% | 13% | 13% |
| Neurological | 3% | - | 7% |
| Percentage taking Pain Medication | 64% | 60% | 67% |
| Anti-Inflammatory (Celebrex)* | 30% | 78% | 20% |
| Pain Killer (Tremadol)* | 79% | 67% | 90% |
| Benzodiazepine (Valium)* | 21% | 44% | 10% |
| Anti-Depressants (Endep)* | 16% | 22% | 10% |
| Engaging in Physiotherapy | 47% | 53% | 40% |
| Past Surgery | 20% | 27% | 13% |
| Other Pain Management (Chiropractor) | 77% | 87% | 67% |
| Depression and Anxiety Disorder | 20% | 20% | 20% |
| Anti-Anxiety Medication* | 33% | 67% | - |

*Note.* CLBP Classification = classification of chronic lower back pain based on Koes *et al* (2001), Non-Specific = no radiographical injury at time of participation, Specific = Radiographical evidence, Neurological = related to change in the Central Nervous System. Pain Average = Average pain intensity one week prior to
participation. Other = Acupuncture, Chiropractor, Massage. \* = Percentage based on individuals taking pain medication

### General Procedure

Demographic and pain-related information were collected via self-report questionnaire. All participants were asked to continue their normal medication routine. Neuropsychological and clinical measures were completed at baseline and immediately following the 4-week brain stimulation. Neuropsychological measures took approximately 2.5 hours to complete.

### Measures

Age, sex, level of education, diagnosis duration, and medication use were collected via self-report questionnaire. All participants completed the neuropsychological and clinical assessment. Outcomes were organised into five functional domains based on theoretical and empirical relationships: Cognition (Executive Function/Attention, Memory, and Language), Pain & Physical Function, and Mood & Wellbeing.

### Neuropsychological Assessment

The neuropsychological measures have been previously reported (see Corti et al., 2021). **Executive Function and Attention** was assessed using the *Stockings of Cambridge* subtest from the Cambridge Neuropsychological Test Automated Battery (SOC; CANTAB™), *Controlled Oral Word Association Task* (COWAT; Benton, Hamsher & Sivan, 1994), *Letter-Number Sequencing* subtest from the Wechsler Adult Intelligence Scale-IV (LNS; Wechsler, 2008), *Stroop (Colour-Word) Test* (Stroop; Jensen & Rohwer, 1966), *Judgement of Line Orientation* (JLO; Benton et al., 1994), and *Hooper Visual Organisation Test* (HVOT; Hooper, 1983). **Memory** was assessed using the *Hopkins Verbal Learning Test-Revised* (HVLT; Brandt, 2001) and *Paragraph Recall* subtest of the Rivermead Behavioural Memory Test (Wilson et al., 1989). **Language** was assessed using the *Boston Naming Test* (BNT; Kaplan et al., 2001) and the *Similarities* subtest of the Wechsler Adult Intelligence Scale-IV (Wechsler, 2008).

### Clinical Assessment

**Pain and Physical Function** was assessed using six outcomes. The *Short-Form McGill Pain Questionnaire* (SF-MPQ) was used to assess pain intensity. The SF-MPQ contains a 10 cm Visual Analogue Scale (VAS; scored 0 – 10) used to assess average pain intensity over the last week (Melzack, 1975). The *RAND 36-Item Short-Form Health Survey* assessed bodily pain (RAND Pain), physical function (RAND Physical Function), and physical limitations (Physical Function; Ware & Sherbourne, 1992). The *Roland-Morris Disability Questionnaire* (RMDQ) assessed the level of disability in CLBP (Roland & Morris, 1983). The *Pain Catastrophising Scale* (PCS) assessed the presence of pain catastrophising (Sullivan et al., 1995).

**Mood and Wellbeing** was assessed using eight outcomes. The *Depression, Anxiety, and Stress Scale-21* (DASS-21) assessed the presence of depression, anxiety, and stress in CLBP (Lovibond & Lovibond, 1995). The *RAND 36-Item Short-Form Health Survey* assessed emotional limitations, fatigue, emotional wellbeing, social functioning, and general health (Ware & Sherbourne, 1992).

### Brain Stimulation

As reported in Corti et al. (2022), participants completed 8-sessions of tDCS stimulation over 4-weeks (2 sessions per week). tDCS was delivered using a constant current stimulator (Soterix®). Participants were randomly assigned (1:1 using block randomisation) to the anodal (a)-tDCS or sham (s)-tDCS group. Participants in the a-tDCS group received 20 minutes of constant 1.5 mA stimulation over left DLPFC every session. The stimulation was administered using two 35 cm2 sponge electrodes soaked in saline solution. The anode electrode was placed over F3 according to the 10-20 international system for EEG electrode placement, to stimulate the left DLPFC. The reference electrode was placed above the left eye, to ensure the current flowed through the prefrontal area. There was a period of 30 seconds at the beginning and end of the tDCS for ramp up/down. Participants in the s-tDCS experienced the 30 second ramp up/down of tDCS but the stimulation was ceased after the 30 seconds. The ramping up of the tDCS aims to provide participants with the initial experience of receiving active tDCS (Ambrus et al., 2012; Nitsche et al., 2008).

### Statistical Analysis

All analyses were conducted using Bayesian multilevel models implemented in R (Version 4.4.3) with the brms package (Bürkner, 2017). A difference-in-differences framework was employed to assess intervention effects, comparing pre-post changes between the a-tDCS and s-tDCS groups. For each outcome, Bayesian linear mixed models were fit with weakly informative priors: normal (0, 1) for regression coefficients, exponential (2) for group-level standard deviations, and exponential (1) for residual variance. Models were estimated using Markov Chain Monte Carlo sampling with 4 chains, 4,000 iterations per chain (1,000 warmup), and convergence was assessed via R^ statistics (all R^ ≤ 1.01). Posterior medians and 95% highest posterior density (HPD) credible intervals are reported. Effect sizes are reported in standard deviation (SD) units. Following individual outcome analyses, an internal meta-analysis was conducted to estimate pooled effects within five predefined domains (Thurston et al., 2009): Cognition: Executive Function/Attention, Cognition: Memory, Cognition: Language, Pain and Physical Function, and Mood & Wellbeing.

## Results

### Individual Outcomes

Table 2 presents the difference-in-differences estimates for all 26 outcome measures. Individual outcomes are organised by domain, with effect sizes representing the differential change (a-tDCS vs. s-tDCS) from pre- to post-intervention. Of the 26 outcomes, only the LNS showed an intervention effect where the HPD did not include 0 (ES = 0.56, 95% HPD [0.09, 1.04]).

**Table 2.** Difference-in-Differences Estimates for All Outcomes by Domain.

| **Domain and outcome** | **Estimate** | **95% HPD CI** |
| --- | --- | --- |
| Cognition: Executive Function/Attention |  |  |
| Letter-Number Sequencing Total | 0.56 | [0.09, 1.04] |
| Stroop Total | 0.33 | [-0.10, 0.75] |
| COWAT Total | 0.19 | [-0.12, 0.50] |
| HVOT Total | 0.11 | [-0.32, 0.50] |
| JLO Total | 0.09 | [-0.43, 0.64] |
| SOC Total | -0.05 | [-0.66, 0.57] |
| **Pooled effect** | **0.21** | **[0.03, 0.40]** |
| Cognition: Memory |  |  |
| HVLT Delayed Recall | 0.17 | [-0.30, 0.63] |
| Paragraph Recall Total | 0.16 | [-0.41, 0.68] |
| Paragraph Delayed Total | 0.08 | [-0.47, 0.63] |
| HVLT Total (3 trials) | -0.05 | [-0.58, 0.46] |
| **Pooled effect** | **0.09** | **[-0.18, 0.35]** |
| Cognition: Language |  |  |
| Similarities Total | 0.32 | [-0.10, 0.77] |
| BNT Total | 0.10 | [-0.27, 0.52] |
| **Pooled effect** | **0.18** | **[-0.11, 0.48]** |
| Pain & Physical Function |  |  |
| RAND Pain | 0.42 | [-0.10, 0.91] |
| Pain VAS | -0.31 | [-0.84, 0.23] |
| Disability | -0.28 | [-0.79, 0.24] |
| Physical Function | 0.18 | [-0.39, 0.77] |
| Physical Limitations | 0.05 | [-0.45, 0.56] |
| Pain Catastrophising | 0.03 | [-0.43, 0.46] |
| **Pooled effect** | **0.18** | **[-0.03, 0.38]** |

| Domain and outcome | Estimate | 95% HPD CI |
| --- | --- | --- |
| Mood & Wellbeing |  |  |
| Stress | -0.38 | [-0.89, 0.12] |
| Social Functioning | 0.28 | [-0.26, 0.82] |
| Depression | -0.09 | [-0.53, 0.34] |
| Anxiety | -0.09 | [-0.66, 0.50] |
| Emotional Wellbeing | -0.03 | [-0.50, 0.44] |
| General Health | -0.10 | [-0.55, 0.36] |
| Fatigue | -0.15 | [-0.54, 0.29] |
| Emotional Limitations | -0.22 | [-0.83, 0.36] |
| **Pooled effect** | **0.03** | **[-0.15, 0.22]** |

*Note.* Values represent difference-in-differences estimates (standardised). For outcomes where higher scores indicate better functioning (RAND measures, cognitive tests), positive values indicate greater improvement in the a-tDCS group. For outcomes where lower scores indicate better functioning (Pain VAS, Disability, Pain Catastrophising, Depression, Stress, Anxiety), negative values indicate greater improvement (symptom reduction) in the a-tDCS group. HPD CI = highest posterior density credible interval. COWAT = Controlled Oral Word Association Test; HVOT = Hooper Visual Organisation Test; JLO = Judgment of Line Orientation; SOC = Stockings of Cambridge; HVLIT = Hopkins Verbal Learning Test; BNT = Boston Naming Test; VAS = Visual Analogue Scale.

### Meta-Analytic Results by Domain

Figure 2 presents individual outcome effects alongside domain-level pooled estimates from the internal meta-analysis. The meta-analytic model revealed heterogeneous effects across cognitive and functional domains.

**Figure 2.**
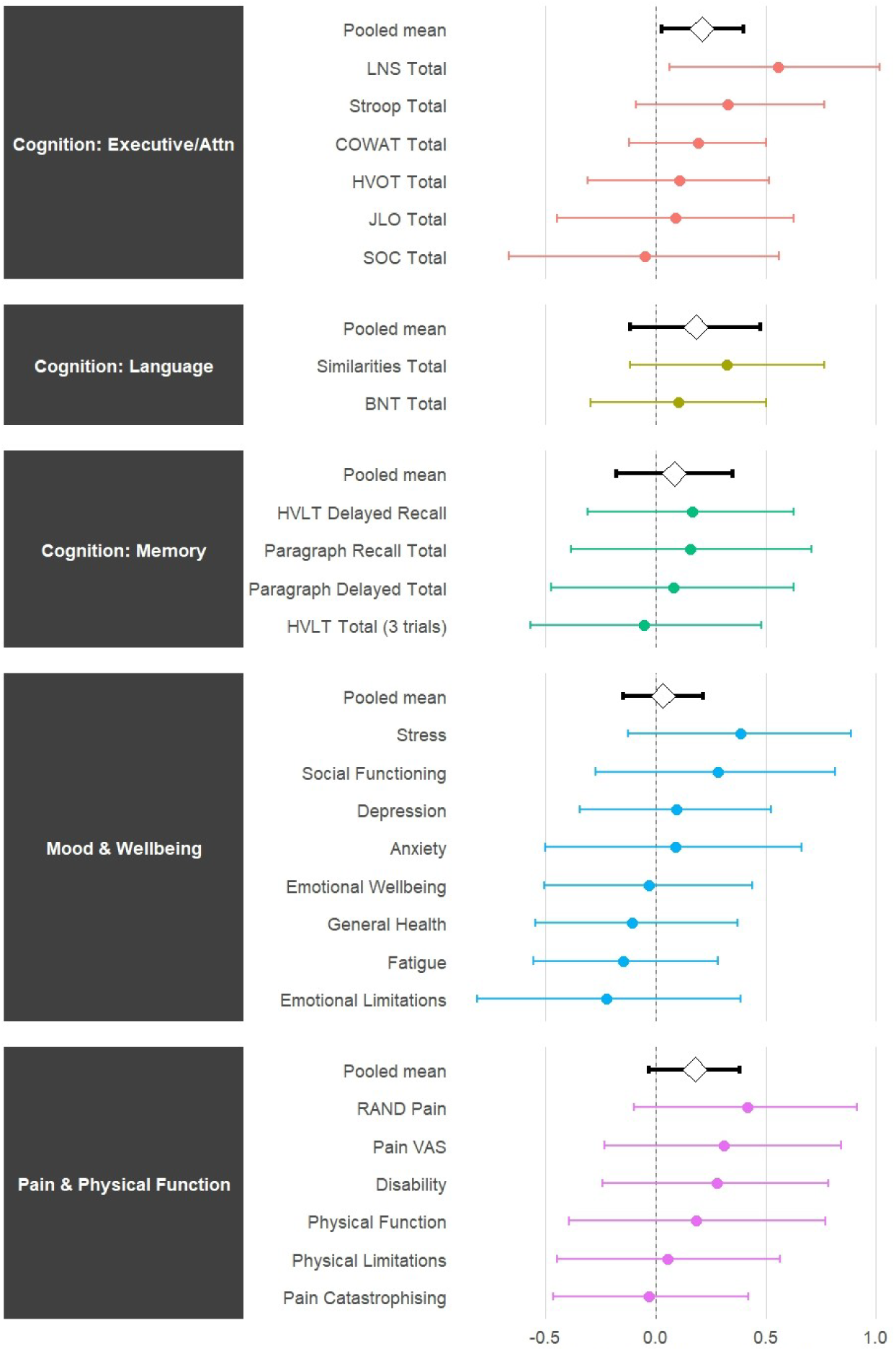
Intervention Effects by Domain with Meta-Analytic Pooled Estimates. *Note.* Points represent difference-in-differences estimates for individual outcomes (Active minus Sham) with 95% highest posterior density credible intervals. Diamonds represent pooled domain-level effects from internal meta-analysis. All effects are direction-aligned a-tDCS group. For measures where lower raw scores indicate better functioning (Pain VAS, Disability, Pain Catastrophising, Depression, Stress, Anxiety), signs were reversed prior to plotting. The vertical dashed line at zero represents no effect. LNS = Letter-Number Sequencing; COWAT = Controlled Oral Word Association Task; HVOT = Hooper Visual Organization Test; JLO = Judgment of Line Orientation; SOC = Stockings of Cambridge; BNT = Boston Naming Test; HVLT = Hopkins Verbal Learning Test; VAS = Visual Analogue Scale.

**Cognition: Executive Function/Attention** demonstrated the most robust evidence of intervention benefit, yielding a pooled effect of 0.21 SD (95% HPD [0.03, 0.40]). The credible interval excluded zero, indicating a reliable domain-level improvement for the a-tDCS group.

**Cognition: Memory** showed a pooled effect of 0.09 SD (95% CI [-0.18, 0.35]). The credible interval included zero, indicating insufficient evidence of a domain-level effect.

**Cognition: Language** showed a pooled effect of 0.18 SD (95% CI [-0.11, 0.48]). While the point estimate suggested a small-to-moderate benefit, the credible interval included zero.

**Pain & Physical Function** yielded a borderline pooled effect of 0.18 SD (95% HPD [-0.03, 0.38]), with the credible interval narrowly overlapping zero. Four of the six individual outcomes (pain intensity, bodily pain, disability, and physical function) showed trends favouring a-tDCS (effect estimates ranging from 0.05 to 0.42 SD).

**Mood & Wellbeing** showed the smallest pooled effect of 0.03 SD (95% CI [-0.15, 0.22]). The credible interval included zero, providing no evidence of intervention effects on emotional wellbeing.

The heterogeneity in domain-level effects (τ² = 0.02–0.04) suggests a-tDCS produced domain-specific rather than generalised effects, with a-tDCS demonstrating selective efficacy for executive cognitive processes while showing limited impact on pain and physical symptoms and negligible effects on emotional wellbeing.

## Discussion

The present study examined if twice weekly, 1.5mA a-tDCS over left DLPFC improved cognitive functioning, pain and disability, and mood and wellbeing in people with CLBP. The findings indicate domain-specific intervention effects. The cognitive domain encompassing Executive Function/Attention demonstrated robust evidence of benefits, with a pooled effect of 0.21 SD (95% HPD [0.03, 0.40]). This effect was driven primarily by improvements in performance on the Letter Number Sequencing task in the a-tDCS group. Pain & Physical Function revealed an uncertain effect that may represent either small analgesic benefits or no effect. In contrast, Mood & Wellbeing showed no evidence of intervention benefit (pooled effect 0.03 SD, 95% HPD [-0.15, 0.22]), with credible intervals for all measures overlapping zero. These findings indicate that a-tDCS over left DLPFC produces selective cognitive benefits in people with CLBP, with uncertain effects on pain and no benefits for mood and emotional wellbeing.

### Cognitive Effects

Executive function and attention performance have been shown to be reduced in individuals with chronic pain (including CLBP) when compared to healthy controls (Berryman et al., 2013; Bunk et al., 2019; Corti et al., 2021; Moore et al., 2019). A-tDCS over left DLPFC has been shown to improve cognitive performance in several populations, including healthy ageing, neurological, and acute and chronic pain conditions (Li et al., 2025; Narmashiri & Akbari; 2025; Silva et al. 2017). In the present study, the Executive Function/Attention domain showed the strongest evidence of a-tDCS benefit, with the credible interval excluding zero and one individual outcome (Letter-Number Sequencing) demonstrating a substantial effect of 0.56 SD (95% HPD [0.09, 1.04]). These effects appear specific to executive processes rather than representing generalised cognitive enhancement, as the Memory and Language domains showed smaller pooled effects (0.09 SD and 0.18 SD respectively). The selective improvement in executive function/attention suggests that a-tDCS over left-DLPFC modulates specific prefrontal cognitive processes rather than producing generalised cognitive improvement.

These findings align with the neuroanatomical targeting of the intervention. The left DLPFC is critically involved in working memory maintenance, attentional control, and executive function (Zgaljardic et al., 2010). A-tDCS over this region has been shown to enhance prefrontal cortex activation and improve top-down cognitive control processes (Deldar et al., 2018; Deldar et al., 2019). The present findings extend this evidence to individuals with CLBP. The magnitude of the effect (0.21 SD at the domain level, 0.56 SD for working memory specifically) suggests a-tDCS over left DLPFC may result in clinically meaningful cognitive improvements in CLBP. In contrast, other cognitive domains showed smaller effects, with credible intervals overlapping zero. This pattern of domain-specific enhancement supports the interpretation that DLPFC stimulation selectively modulates circuits underlying executive control and attention, rather than producing non-specific cognitive facilitation.

### Pain and Physical Function Effects

The Pain & Physical Function domain demonstrated an uncertain effect (0.18 SD, 95% HPD [-0.03, 0.38]). While the credible interval marginally overlaps zero, four of six measures in this domain showed numerical trends favouring a-tDCS, including pain intensity (McGill Pain VAS -0.31, disability (-0.28), bodily pain (RAND Pain 0.42), physical function (Physical Function 0.18). The PCS and Physical Limitations showed negligible change (0.03 and 0.05 SD, respectively). The consistent directional trends across multiple pain and physical function measures suggest that if a-tDCS over left DLPFC produces analgesic and functional benefits in CLBP, they are likely small in magnitude.

The small pain effects observed in the present study are consistent with the limited and heterogeneous literature on a-tDCS over DLPFC for chronic pain. While early small-scale studies suggested analgesic benefits of DLPFC stimulation, particularly in fibromyalgia (Fregni et al., 2006), more recent trials have reported null or inconsistent effects (Samartin-Veiga et al., 2022). Meta-analytic evidence classifies a-tDCS over DLPFC for pain as having “possible but uncertain efficacy” (Lefaucheur et al., 2017), reflecting substantial heterogeneity across studies and pain conditions. Meta-analytic evidence indicates that when DLPFC effects on pain are significant, they are typically small in magnitude and less consistent than effects produced by motor cortex (M1) stimulation. Previous research has shown that a-tDCS over M1 produces consistent analgesic effects via modulation of thalamo-cortical pathways in chronic pain populations, including CLBP (Chaturvedi et al., 2022; Pacheco-Barrios et al., 2020). DLPFC stimulation, however, relies on indirect cognitive-affective pain modulation mechanisms. The uncertain pain effects observed in the present study may reflect genuine small effects of left DLPFC stimulation on pain processing or may indicate that this tDCS montage is not optimally positioned to modulate pain networks in CLBP. As such, pain relief in CLBP may require stronger modulation of pain processing networks or may benefit more from alternative montages targeting primary motor or sensory cortices. Future trials are needed to establish whether a-tDCS over DLPFC produces clinically meaningful pain relief and directly compare different montages (e.g., M1 vs DLPFC) to determine which cortical targets are more effective for pain modulation in CLBP.

### Mood and Wellbeing Effects

The Mood & Wellbeing domain demonstrated the smallest pooled effect (0.03 SD, 95% HPD [-0.15, 0.22]), with credible intervals for all eight measures substantially overlapping zero. This domain included measures of depression, anxiety, stress, fatigue, emotional wellbeing, social functioning, and general health. The near-zero pooled effect and lack of consistent directional trends across measures suggests that a-tDCS over left-DLPFC does not improve psychological distress or emotional wellbeing in people with CLBP.

The absence of mood and emotional wellbeing benefits has important clinical implications for individuals with CLBP. Many individuals with CLBP report higher levels of depression, anxiety, and reduced quality of life compared to age and gender-matched controls (Corti et al., 2021). The present findings indicate that a-tDCS over left DLPFC is insufficient to produce large, meaningful improvements in these psychological dimensions of CLBP. This suggests that adjunctive psychological interventions targeting mood and emotional wellbeing remain necessary for comprehensive CLBP management.

### Mechanisms Underlying Domain Specificity

The pattern of heterogeneous effects across the domains has implications for understanding tDCS mechanisms in CLBP. Three distinct patterns emerged: robust effects for executive function/attention, small uncertain effects for pain, and null effects for mood and wellbeing. The domain specificity indicates that a-tDCS over left-DLPFC produces targeted neuromodulation rather than generalised, non-specific effects.

The dissociation between cognitive and pain outcomes could be explained via several mechanisms. First, left DLPFC may be optimally positioned to modulate executive control circuits, while being less effective at modulating pain processing networks that involve motor, sensory, and subcortical structures (Seminowicz & Moayedi, 2017). Secondly, the stimulation procedure (1.5mA, twice weekly, four weeks) may be adequate for modulating cognitive function but insufficient for pain relief, which may require higher intensity or more frequent stimulation. While the optimal tDCS protocol for pain reduction in chronic pain is still under investigation, research in fibromyalgia reported a median of 15 stimulation sessions of high definition-TDCS is required to reduce perceived pain by at least 50% (Castillo-Saavedra et al., 2016). Individual differences in pain mechanisms may also moderate treatment response, such that left DLPFC stimulation only benefits a subset of individuals with CLBP with specific cognitive-affective pain profiles (Zortea et al., 2019).

As executive function/attention improvements occurred without corresponding changes in pain or mood, this suggests that improved cognition does not automatically translate to improved (i.e., reduced) pain or emotional wellbeing. This challenges simplistic models whereby improved executive function/attention would enhance pain coping or mood regulation (Berginström et al., 2024; Bunk et al., 2019). As such, the absence of mood and wellbeing benefits may reflect distinct neural and physiological mechanisms underlying mood disturbances in CLBP. While DLPFC stimulation effectively modulates executive control circuits, mood disturbances in CLBP may involve different neural systems, such as limbic and corticostriatal reward circuits (Baliki et al., 2012), as well as systemic factors such as inflammatory processes (Burke et al., 2017; Miller & Raison, 2016). Additionally, mood is often reactive to ongoing pain experiences and maintained by pain catastrophising (Sullivan et al., 2001). The present study found negligible changes in pain catastrophising, which may explain the absence of mood improvements, as catastrophising has been found to mediate the relationship between pain and emotional distress (Edwards et al., 2006; Quartana et al., 2009). Furthermore, DLPFC may not be optimally positioned to modulate mood regulation circuits, which involve ventromedial prefrontal cortex and anterior cingulate regions (Etkin et al., 2011). This suggests that effective mood interventions in CLBP may require either targeting different cortical regions or combining brain stimulation with cognitive-behavioural approaches that directly address catastrophising and maladaptive pain beliefs.

### Limitations

Several limitations warrant consideration. The use of analgesic medications can influence cognitive functioning, pain, and psychological outcomes. While medication use was recorded in the present study, frequency of use and dosage was not documented. It is unknown if medication use was similar, reduced, or increased between pre- and post-intervention. We cannot rule out if changes in analgesic medication use may play some role in the observed improvements in pain-related and psychological outcomes. Previous research has produced conflicting results as to whether the use of analgesic medication hinders or improves cognitive future (Higgins et al., 2018). Future research should consider the inclusion of non-medicated participants to reduce any potential influence of medication on cognitive function. However, this may be difficult to achieve given that chronic pain is usually managed through medication.

The design of the study makes it difficult to differentiate between the effect of tDCS and the effect of engaging in the intervention. Previous research has reported trial characteristics (e.g., the number of visits) were significantly associated with increased placebo response (Tuttle et al., 2015; Vase et al., 2015). Participants in the present study attended 9 appointments over a 5-week period, and it is possible that this may have impacted on both physical and emotional health. Attending the appointments may have increased participants movement and physical functioning and may have increased perceived social support (from the researchers). These factors would be expected to affect both groups similarly and cannot explain domain-specific between-group differences, however, they may have contributed to improvements observed in both groups across multiple outcomes. Future research should also consider including a no-intervention group, who receive the same level of social support as the intervention group, to better isolate the specific effects of tDCS from non-specific study-related factors.

### Conclusion

The present findings suggest that a-tDCS over left DLPFC produces domain-specific effects in CLBP, with improvements in executive function/attention, uncertain analgesic and functional effects, and no meaningful benefit on mood and emotional wellbeing. The pattern of domain-specific effects indicates targeted neuromodulation of prefrontal cognitive circuits rather than generalised, non-specific improvements. These findings establish a-tDCS over left DLPFC as a potentially useful intervention for addressing cognitive deficits in CLBP but is unlikely to serve as an effective standalone analgesic or mood intervention. The dissociation between cognitive, pain, and emotional outcomes highlights the need for further investigation of the potential clinical benefit of combining tDCS with adjunctive psychological interventions targeting mood and emotional wellbeing in CLBP.

## Data Availability

All data produced in the present study are available upon reasonable request to the authors

